# The prevalence of post COVID-19 condition (PCC) and a simple risk scoring tool for PCC screening on Bonaire, Caribbean Netherlands: a retrospective cohort study

**DOI:** 10.1101/2023.06.20.23291646

**Authors:** Danytza SF Berry, Thomas Dalhuisen, Giramin Marchena, Ivo Tiemessen, Eveline Geubbels, Loes Jaspers

## Abstract

**Aim:** To assess the prevalence of post COVID-19 condition (PCC) on Bonaire and develop a practical risk scoring tool for PCC screening, using easily obtainable characteristics.

**Methods:** A retrospective cohort study of symptomatic SARS-CoV-2 cases were randomly sampled from Bonaire’s case-registry and telephone interviewed between 15-November-2021 and 4-December-2021. PCC patients had a PCR-positive SARS-CoV-2 test (1-March-2020 and 1-October-2021) and self-attributed at least one symptom lasting over four weeks to their infection. Multivariate logistic regression was used to derive a risk formula to develop a practical risk scoring tool.

**Results:** Out of 414 cases, 160 (39%) were PCC patients. Fifty-three patients were unrecovered (median illness duration 250 days (IQR 34)). Of recovered patients, 35% experienced symptoms for at least 3 months after disease onset. PCC prevalence was highest among females (38%), 40-59 year-olds (40%), morbidly obese (31%) and hospitalized patients (80%). A PCC risk scoring tool using age, sex, presence of comorbidities, and acute phase hospitalization or GP visit had an area-under-the-curve (AUC) of 0.68 (95%CI 0.63-0.74). Adding smoking, alcohol use, BMI, education level, and number of acute phase symptoms increased the AUC to 0.79 (95%CI 0.74-0.83). Subgroup analyses of non-hospitalized patients (n=362) resulted in similar AUCs.

**Conclusion:** Thee estimated prevalence of PCC on Bonaire was 39%. Moreover, easily obtainable patient characteristics can be used to build a risk scoring tool for PCC with acceptable discriminatory power. After external validation, this tool could aid the development of healthcare interventions in low resource settings to identify patients at risk for PCC.

**Key messages:** *What is already known on this topic:*

- An increasing number of studies show that varying proportions of COVID-19 patients are left with prolonged health issues, including persistence of symptoms such as fatigue, shortness of breath, loss of muscle strength, and concentration issues following the acute phase of COVID-19.
- Prevalence of this recently recognized medical syndrome, ‘long-COVID’ or ‘post COVID-19 condition (PCC)’, ranges between 37-49% in the European Netherlands, though there are no reports of PCC prevalence in the Caribbean Netherlands to date.

*What this study adds:*

- We designed the first retrospective cohort study estimating the prevalence and characteristics of PCC on Bonaire, Caribbean Netherlands and devised a simple risk scoring formula to calculate PCC risk.
- We found a PCC prevalence of 39% on Bonaire and describe a proof-of-concept risk scoring tool with good discriminatory performance.

*How this study might affect research, practice or policy:*

- Our study estimates PCC prevalence and describes disease and patient characteristics for Bonaire. This is the first study providing these insights in the Caribbean; a region that has been underrepresented in PCC research.
- Furthermore, our study highlights the added benefit of registering sufficient patient characteristics at the time of consultation for acute COVID-19 disease, for improved PCC screening later on.
- After external validation of our proof-of-concept study, this risk scoring tool could aid the development of primary care and public health interventions and health communication strategies in low resource settings for the identification of patients at risk for PCC.

## INTRODUCTION

As of 11 January 2023, an estimated 660,378,145 confirmed COVID-19 cases including 6,691,495 deaths have been reported worldwide [1]. Although most cases fully recover shortly after the acute infection, an increasing number of international studies suggest that varying proportions of patients are left with prolonged health issues [2]. This recently recognized medical syndrome, ‘long-COVID’ or ‘post COVID-19 condition (PCC)’, includes persistence of symptoms like fatigue, shortness of breath, loss of muscle strength, and concentration issues following the acute phase of COVID-19 [3–5]; much like other post-infectious syndromes, where prolonged symptoms can occur in a remitting pattern due to persistence of an infectious agent, such as Lyme disease or Q-fever [2, 5].

In van der Maaden et al. (2023), a longitudinal study of PCC in the European Netherlands, 49% of COVID-19 cases reported symptoms three months after SARS-CoV-2 infection [6]. A study by Nivel showed 37% of Dutch COVID-19 patients visiting the GP three months after disease onset reported still experiencing at least one symptom [7]. These findings are roughly in line with a recent global pooled PCC prevalence estimate of 43% (95% CI, 39-46%), though untangling symptom attribution solely to SARS-CoV-2 infection remains challenging [2]. Furthermore, PCC prevalence appears higher among those hospitalized (54%, 95% confidence interval (CI) 44-63%) compared to non-hospitalized patients (34%, 95% CI 24-46%) [2].

Since the start of the COVID-19 pandemic, Bonaire has seen various waves of SARS-CoV-2 infections. Bonaire is an island in the Dutch Caribbean with 21,745 permanent residents in 2021 [8]. By 1 October 2021, 2,036 persons living or staying on the island had tested positive, of whom 57 were admitted to the hospital and 19 passed away due to or with COVID-19 [9]. In May 2021, local general practitioners (GPs) on the island raised concerns about the lack of knowledge on the burden and risk factors of PCC among Caribbean populations and the impact of PCC on health systems in Caribbean settings. Local practitioners hypothesized the prevalence and symptomatology of PCC may differ substantially from that reported in Western settings due to the different health system context, demographics, and burden of non-communicable disease (NCD) on Bonaire [10–12]. Various studies have shown disparities between different populations in COVID-19 risk and severity [13], though little remains known of how these disparities translate into PCC outcomes and care [14, 15].

Besides gaining insights into the prevalence of PCC on Bonaire, practical and easy-to-use risk scoring tools are needed to identify cases in low resource-settings that are at an increased risk of developing PCC. Formal PCC diagnostics are lacking, and PCC consequently remains an underdiagnosed condition even though timely diagnosis is needed to access appropriate insured care and treatment [15, 16]. Tools that may identify those at an increased PCC risk can aid in increasing the proportion of PCC patients that are formally diagnosed by healthcare providers in a timely matter. Ideally, these tools should require only a limited number of inputs, which can be easily obtained by GPs. Therefore, in this study we aim to assess the prevalence and characteristics of PCC on Bonaire and to develop a pragmatic risk scoring tool using easily obtainable patient characteristics to identify patients at risk of developing PCC.

## METHODS

### Study design & setting

We designed a retrospective cohort study of individuals who tested positive for SARS-CoV-2 at the public health department of Bonaire between the start of the pandemic and 1 October 2021 (**Supplementary Figure 1**). The public health department was the main facility on the island for community testing. Testing was free of charge. The type of diagnostic tests used were in accordance with the guidelines from the Dutch Ministry of Health at the time of testing and included approved PCR- or antigen tests. The study protocol was developed through a collaboration between researchers from the Public Health Department on Bonaire and the National Institute for Public Health and the Environment (RIVM, in European Netherlands). Data collection took place on Bonaire, and data analysis was carried out by researchers from the RIVM in close virtual collaboration with the researchers based on Bonaire.

### Participants

Participant data was exported from HP Zone, the patient registration and outbreak management software system used to register data about the notifiable cases throughout the COVID-19 pandemic on Bonaire. Individuals were eligible for recruitment if they were a) permanent residents of Bonaire; b) symptomatic; c) had a laboratory confirmed SARS-CoV-2 positive test result, and d) had contact details available at the public health department. All eligible participants were assigned a number: 0 for patients hospitalized for COVID-19 during the acute phase or randomly the number 1, 2, 3 or 4 for non-hospitalized patients. All patients hospitalized for COVID-19 during the acute phase were invited for study participation. Thereafter, all the eligible participants in groups 1-4 were called, starting with number 1, until enough participants were included to satisfy the required power threshold of the study. This approach allowed us to incorporate a random element of sample selection and reaching enough power for the study by calling all patients hospitalized for COVID-19 during the acute phase first. This approach was chosen, given that on Bonaire, resources for the study completion were minimal, and we wanted to assure maximum power for the study.

### Patient and public involvement

Local GPs were provided the opportunity to help design the questionnaire and to formulate research priorities. Preliminary results were shared with the public by Bonaire’s Public Health Department through local media (radio and television) and a press briefing in the spring of 2022. Moreover, the results were discussed in a group session of health care and governmental professionals, including representatives of the GPs, the hospital, the health insurance office, and policy advisors of the local government. Remaining research needs were addressed, and where possible, integrated into the ongoing analyses.

Furthermore, COVID-19 patients were able to self-enroll in the study if they were not called by the interviewers, but felt the need to share their experiences. They were given a separate coding in the questionnaire, in order to distinguish the randomly selected from the self-selected participants. In the end, no patients opted to self-enroll in the study.

### Case definitions and community controls

A PCC patient was defined as “an individual with a laboratory confirmed SARS-CoV-2 positive test result, of whom at least one symptom self-attributed to the experienced SARS-CoV-2 infection lasted longer than four weeks” [17]. Cases with a laboratory confirmed SARS-CoV-2 positive test result, of whom all symptoms self-attributed to the experienced SARS-CoV-2 infection lasted shorter than four weeks were defined as non-PCC cases. A community control group was included to assess background prevalence of symptoms. By doing so, we aimed to have a stronger basis to differentiate whether certain results could be attributable to pre-existing underling disease, to a pandemic effect, or to the experienced SARS-CoV-2 infection. During the telephone interview, SARS-CoV-2 positive cases were asked to provide contact details of a community control, defined as “an individual who had not been a close contact of the case during the infection phase, was of the same sex and similar age, was from outside their household, and who they knew had not tested positive with SARS-CoV-2 until the moment of interview”. It was expected that this approach would result in more controls as opposed to recruiting individuals who had tested negative at the island’s testing facilities.

### Study size

The expected study size was calculated based on a desired alpha of 0.05, power of 80%, expected response rate of 70% and a conservatively estimated expected PCC prevalence of 20% at 4 weeks after acute infection. Therefore, a sample of roughly 600 COVID-19 patients would be needed to estimate the expected 20% prevalence of PCC with a 95% confidence interval of 6% width [17-23%]. In addition, a cohort of 200 self-reported SARS-CoV-2 negative individuals was invited in order to be able to compare the distribution of risk factors for and symptoms of PCC among COVID-19 patients with that of the general population. In total, we aimed to invite 800 individuals to participate in the study.

### Data collection

A questionnaire was designed based on 2021 PCC surveys from Nivel (Netherlands Institute for Health Services Research), RIVM (Netherlands National Institute for Public Health and the Environment), GGD Zuid Holland Zuid (Municipal Health Service South Holland), Radboud University Medical Centre (UMC), and Maastricht & Hasselt University. Our questionnaire was developed for three groups: those who did not have COVID-19 (community controls), those who did get infected with SARS-CoV-2 but did not develop PCC (non-PCC cases), and those who developed PCC (PCC patients).

Survey questions focused on demographics, health status prior to the first outbreak, pre- and post-infection COVID-19 vaccination status, hospitalization for acute COVID-19 symptoms, PCC symptoms at the time of the interview, and healthcare utilization pre- and post-infection. The questionnaire was reviewed for practical relevance by several local GPs. It was translated into the four main languages spoken on Bonaire: Dutch, English, Papiamentu, and Spanish. Thereafter, the questionnaires were uploaded into an online form using the data management program ‘digitale checklisten.nl’ [18]. It was expected this approach would increase the response rate, taking local health literacy into consideration. No back translation procedure took place due to time constraints.

In collaboration with the employment agency Tempo [19], we developed a job vacancy and hired a team of 30 local interviewers. In November 2021, they were trained to carry out telephone interviews by the research team of the Public Health Department and the Central Bureau of Statistics (CBS) Bonaire.

Eligible respondents were invited to participate in the study through a WhatsApp message, which included information about the study and the informed consent statement (both in writing and as a voice message recorded by a well-known Bonairean spokesperson). Verbal informed consent was obtained and documented by the main interviewer prior to the start of the interview. Respondents were explained the purpose of the study and that they could choose to resign from the interview at any time. A second interviewer verified whether consent was given, prior to the first interviewer commencing the interview. The second interviewer only confirmed whether the informed consent was given by the respondent. Data was collected between 15 November and 4 December 2021 through telephone interviews of approximately 20 minutes for community controls, 30 minutes for non-PCC cases, and 45 minutes for PCC-patients. Eligible respondents were called inside and outside office hours to increase the likelihood of being reached. Interviewers made three attempts to reach an eligible respondent, otherwise they were registered as not reached on the call list. Data was directly entered into the online registry system ‘digitale checklisten’ by the interviewer.

### Ethical approval

The study protocol was sent to the Central Committee on Research Involving Human Subjects (CCMO Netherlands) [20], who confirmed on 8 October 2021 that the study did not require ethical approval due to its observational design. Patient data export necessary for participant selection was only extracted from HP zone after permission from the Dutch COVID-19 patient data protection unit at GGD GHOR – the Netherlands was given.

### Variables

For this study, we defined PCC as “the presence of at least one of the following 14 symptoms minimally 4 after weeks of receiving a positive test result for SARS-CoV-2”, based on the Centers for Disease Control (CDC) their staging at the time: chest pain, concentration problems, cough, fatigue, headache, heart palpitations, loss of appetite, reduced muscle strength, loss of sense of smell, loss of sense of taste, muscle ache, reduced physical endurance, shortness of breath, and sleeping problems [5,17].

Our second objective was to develop a pragmatic risk score formula using variables that could be easily obtained by a GP in clinical practice on Bonaire. We selected predictors with the perspective of the data collection being feasible in GPs’ everyday practice. Hence, these included factors and data that GPs register in the patient file with regard to the COVID-19 infection consultation, factors that are generally available in the GP’s electronic patient database, or factors which are easily accessible upon verbal inquiry with the patient.

Demographic factors included age, sex (male/female), and education level. We categorized age into groups 0-19 years, 20-39 years, 40-59 years, and 60+ years old. Education level was categorized as low, moderate or high [21]. Low education indicated no education, primary level education, lower vocational and lower secondary education; moderate indicated higher secondary education, and high indicated university level post-secondary education. Health-related factors included pre-pandemic smoking (yes/no), alcohol use (yes/no), BMI, and comorbidities. For BMI, we removed one outlier (>70), and categorized remaining values into underweight (<18), normal (≥18, <25), overweight (≥25-<30), obese (≥30-<35) and morbidly obese (≥35).

Comorbidity was categorized as the presence of at least one underlying disease (yes/no). Comorbidities included specific underlying diseases with a diagnosis, such as diabetes, cardiovascular disease, depression, neurocognitive disorders, stroke, and more. COVID-19 related factors included acute phase GP visit, hospitalized for COVID-19 during the acute phase (not stratified by ICU or general care) and number of symptoms present during acute phase. Definitions of variables considered in the analyses are further specified in **Supplementary Table 1**.

### Statistical methods

First, descriptive statistics and univariate analyses were conducted comparing PCC to non-PCC cases and comparing the COVID-19 cohort to community controls using Fisher’s exact test or Wilcoxon test where appropriate. We considered a p-value <0.05 as statistically significant. All analyses were carried out in R version 4.1.3 (R Foundation for Statistical Computing).

#### Sparse and rich models

Thereafter, we developed two sets of models using variables that could be easily obtained by a GP in clinical practice on Bonaire. The first set is sparse and includes patient characteristics that, based on discussions with a local GP, are easily attainable from a GP practice patient file. The variables included in this model are age, sex, presence of one or more comorbidities, hospitalization for COVID-19 during the acute phase and GP visit during the acute phase of a SARS-CoV-2 infection. The second set is richer and includes additional variables not routinely registered in GP patient files but which should be obtainable with relative ease upon a patient’s visit or verbal inquiry by telephone: BMI, lifestyle factors (such as whether a patient smokes or uses alcohol) and the number of symptoms experienced during the acute phase of their SARS-CoV-2 infection. Because having been hospitalized for acute COVID-19 was shown to be a strong predictor of PCC during explorative analyses, within each set we built one model for the full patient sample (hospitalized and non-hospitalized during acute COVID-19) and one model for non-hospitalized patients only. Thus, four models were built:

Models based on the full patient sample including both hospitalized and non-hospitalized patients (n=390) included the following variables:

- Model 1a (sparse model): age, sex, presence of one or more comorbidities, acute phase GP visit, and acute phase hospitalization;
- Model 2a (rich model): age, sex, presence of one or more comorbidities, BMI, smoking status, alcohol use, education level, number of symptoms experienced during acute phase, acute phase GP visit, and acute phase hospitalization.

Models based on the non-hospitalized patient sample (n=362) included the following variables:

- Model 1b (sparse model): age, sex, presence of one or more comorbidities, and acute phase GP visit;
- Model 2b (rich model): age, sex, presence of one or more comorbidities, BMI, smoking, alcohol use, education level, number of symptoms experienced during acute phase, and acute phase GP visit.

#### Development of risk score formula

Multivariate logistic regression models were fit using PCC status as the outcome variable. Regression coefficients from fitted models were then multiplied by 10 and rounded to the nearest integer to derive a risk score formula (**Supplementary Tables 2 and 3**). The risk score formula was then applied to calculate a risk score for each COVID-19 patient. The discriminatory performance of the risk score was then assessed by plotting the receiver operating curve (ROC) and calculating the area under the curve (AUC) and its 95% CI using 2000 stratified bootstrap replicates implemented in the pROC package in R [22]. Discriminatory performance relates to how well the risk score can distinguish between PCC patients and non-PCC cases. We then assessed the risk score formula’s calibration and performed internal validation using the optimism bootstrapping implemented in the rms R package (**Supplementary Figure 2**) [23]. Using the sensitivity and specificity values for each risk score cut-off in the ROC curves, we then calculated the positive predictive value (PPV) and negative predictive value (NPV) of our risk score formula for each risk score cut-off. The PPV and NPV were calculated with the formulas below using this study’s PCC prevalence estimate of 39%:

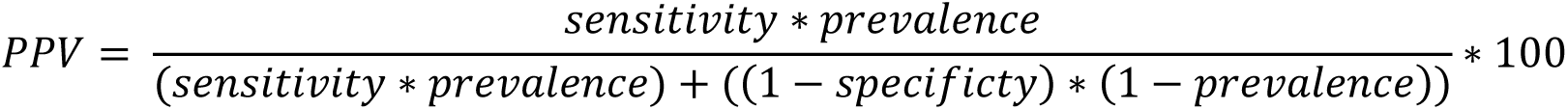

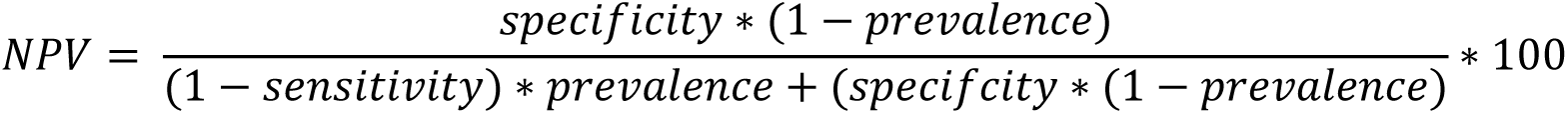

#### Missing data

Prior to fitting multivariate logistic regression models, we examined the extent of missing data for all variables we aimed to include. We opted for a complete case-analysis, thereby excluding n=24 (5.8%) cases from the COVID-19 cohort. Of excluded cases, we confirmed they did not have disproportional baseline patient characteristics.

#### Sensitivity analysis

We investigated whether an intermediate risk scoring tool could attain a similar AUC as our rich risk scoring tool. This intermediate scoring tool uses fewer patient characteristics than our rich risk scoring tool, but more than our sparse risk scoring tool. The variables included were: age, sex, presence of one or more comorbidities, GP visit during the acute phase, alcohol use, smoking, and BMI (**Supplementary Tables 4 and 5**).

## RESULTS

### Participants

All cases recorded in the national case registry ‘HP Zone’ by 1 October 2021 (n=2,036) were assessed for eligibility. Based on the inclusion criteria, 1,789 individuals were eligible for study participation. Of the excluded persons,133 individuals were non-residents, 95 were asymptomatic and 19 had passed away at the time of inclusion. Of the eligible individuals, 357 were not reached after three attempts, 76 did not have time to participate, 66 did not want to participate, 47 had switched phone numbers and were not reachable anymore, 15 did not see a value in participating, four did not understand or trust the interview procedure, and 73 had another, unspecified, reason not to participate.

Of another 545 individuals, reason of non-participation was not recorded and 103 did not give informed consent. Of interviewed individuals, 8 did not finish the interview, three changed their mind about participating, one did not want to clarify whether they had had COVID-19, and of two, date of birth was not registered. In total, 489 respondents were included in the study (**Figure 1**), of whom 414 were lab-confirmed SARS-CoV-2 positive cases and 75 had a self-reported SARS-CoV-2 negative status (median age 43, IQR 26).

**Fig. 1.**
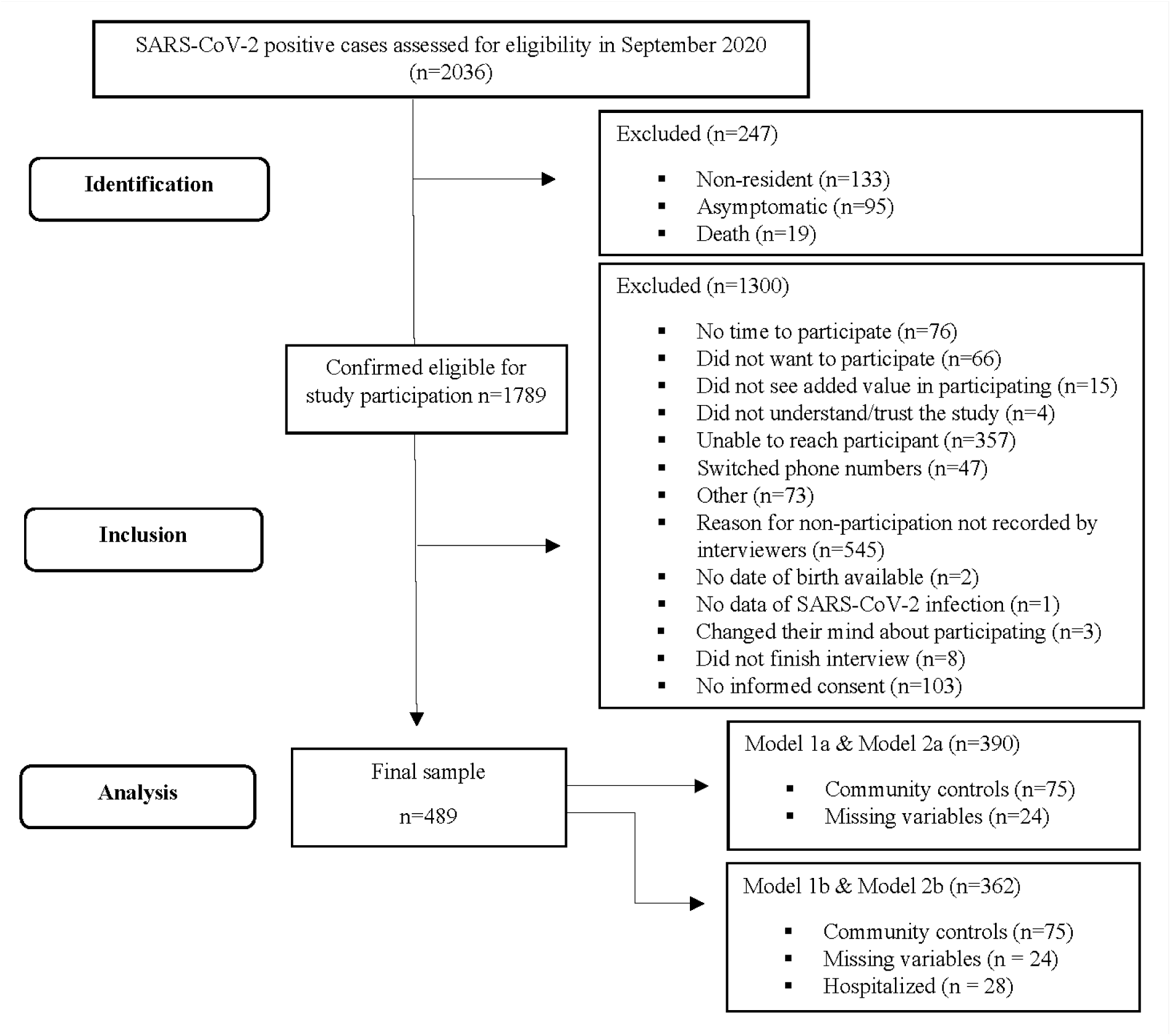
STROBE flowchart describing participant selection for the study examining prevalence and predictive factors of PCC on Bonaire, Caribbean Netherlands

Out of a total of 2,036 SARS-CoV-2 positive cases detected until October 1^st^, 2021, 133 cases were not eligible for inclusion because they were not a permanent resident of Bonaire, 95 because they were asymptomatic, and 19 were not eligible due to non-survival by 1 October 2021. Thus, there were 1,789 cases confirmed eligible for inclusion. Of these, 57 cases had been hospitalized during the acute phase of infection. A total of 1,195 eligible participants were called during the interview period (67%), of whom 986 were reached (83%). The final sample consisted of 503 participants who had given informed consent (52%). Of these, 14 were excluded as they could not be categorized into one of the subgroups, for reasons such as missing date of onset or date of birth or choosing to stop participation during the interview. This flowchart has also been published in the sub-study by Berry et al. (2023) [‘manuscript in preparation’]

### Baseline characteristics

160 cases (39%) fit the case definition of PCC (median age 43, IQR 21). At the time of interview, 107 patients had recovered from PCC. Of recovered patients, 42 (39%) recovered within 2 months after disease onset, 27 (25%) recovered within 3 months, 16 (15%) within 4 months and 21 (20%) patients had not recovered after 4 months of disease onset. Time to recovery was unknown for 1 patient. 53 patients were unrecovered at the time of interview and had a median duration of illness of 250 days (IQR 34).

In all three groups, there were more female participants than males (**Table 1**). PCC prevalence was higher among females (38%) than males (25%), highest in the age group 40-59 years old (40%), highest among morbidly obese patients (31%), and higher in cases hospitalized during the acute phase of infection (80%) as compared to the non-hospitalized group (35%).

**Table 1.** Baseline demographics and univariate analyses to assess between group differences

|  | **PCC patients**<br>**(n=160)** | **Non-PCC cases**<br>**(n=254)** | **Overall positive cohort**<br>**(n=414)** | **Community controls**<br>**(n=75)** | **P value**<br>**(PCC vs. non-PCC cases)** | **P value**<br>**(Community controls vs. overall positive cohort)** |
| --- | --- | --- | --- | --- | --- | --- |
| **Characteristics** |  |  |  |  |  |  |
| **Median age (IQR)** | 43 (21) | 39 (23) | 40 (23) | 43 (26) | **<0.007<sup>A</sup>** | 0.09<sup>A</sup> |
| **Age groups** |  |  |  |  | **<0.003<sup>B</sup>** | 0.14<sup>B</sup> |
| 0-19 | 6 (4%) | 36 (14%) | 42 (10%) | 3 (4%) |  |  |
| 20-39 | 61 (38%) | 98 (39%) | 159 (38%) | 28 (37%) |  |  |
| 40-59 | 64 (40%) | 85 (34%) | 149 (36%) | 25 (33%) |  |  |
| 60+ | 29 (18%) | 34 (13%) | 63 (15%) | 19 (25%) |  |  |
| Unknown | 0 | 1 (0%) | 1 (0%) | 0 |  |  |
| **Sex** |  |  |  |  | **<0.001<sup>B</sup>** | 0.47<sup>B</sup> |
| Male | 46 (29%) | 115 (45%) | 161 (39%) | 25 (33%) |  |  |
| Female | 114 (71%) | 138 (54%) | 252 (61%) | 50 (67%) |  |  |
| Non-binary | 0 | 1 (0%) | 1 (0%) | 0 |  |  |

| Education level |  |  |  |  | 0.15<sup>B</sup> | 0.78<sup>B</sup> |
| --- | --- | --- | --- | --- | --- | --- |
| Low | 119 (74%) | 208 (82%) | 327 (79%) | 57 (76%) |  |  |
| Moderate | 23 (14%) | 29 (11%) | 52 (13%) | 12 (16%) |  |  |
| High | 17 (11%) | 14 (6%) | 31 (8%) | 6 (8%) |  |  |
| Unknown | 1 (1%) | 3 (1%) | 4 (1%) | 0 |  |  |
| **Pre-pandemic factors** |  |  |  |  |  |  |
| Median BMI (IQR) | 30.2 (11.3) | 28.1 (8.71) | 29.0 (9.38) | 28.7 (8.32) | <0.001<sup>A</sup> | 0.78<sup>A</sup> |
| BMI category |  |  |  |  | <0.001<sup>B</sup> | 0.92<sup>B</sup> |
| Underweight | 2 (13%) | 4 (2%) | 6 (1%) | 1 (1%) |  |  |
| Normal | 32 (20%) | 80 (32%) | 112 (27%) | 19 (25%) |  |  |
| Overweight | 41 (26%) | 73 (29%) | 114 (28%) | 22 (29%) |  |  |
| Obese | 36 (23%) | 53 (21%) | 89 (21%) | 19 (25%) |  |  |
| Morbidly obese | 49 (31%) | 42 (17%) | 91 (22%) | 1 (1%) |  |  |
| Unknown | 0 | 2 (1%) | 2 (0%) | 0 |  |  |
| Comorbidities |  |  |  |  | 0.05<sup>B</sup> | 0.40<sup>B</sup> |
| Yes, one or more | 33 (21%) | 33 (13.0 %) | 66 (16%) | 15 (0%) |  |  |
| No | 127 (79%) | 221 (87.0%) | 348 (84%) | 60 (80%) |  |  |
| Smoking |  |  |  |  | 0.45<sup>B</sup> | 0.66<sup>B</sup> |
| Yes | 18 (11%) | 36 (14%) | 54 (13%) | 14 (19%) |  |  |
| No | 142 (89%) | 215 (85%) | 357 (86%) | 61 (81%) |  |  |
| Unknown | 0 | 3 (1%) | 3 (1%) | 0 |  |  |
| Alcohol use |  |  |  |  | 0.29<sup>B</sup> | 0.73<sup>B</sup> |
| Yes | 109 (68%) | 158 (62%) | 267 (65%) | 44 (59%) |  |  |
| No | 51 (32%) | 94 (37%) | 145 (35%) | 30 (40%) |  |  |
| Unknown | 0 | 2 (1%) | 2 (0.5%) | 1 (1%) |  |  |
| **Factors during acute COVID-19** |  |  |  |  |  |  |
| Median number of symptoms | 10 (5) | 5 (7) | 7 (8) | - | <0.001<sup>A</sup> | - |

| present (IQR) |  |  |  |  |  |  |
| --- | --- | --- | --- | --- | --- | --- |
| **GP visit** |  |  |  |  | **0.02<sup>B</sup>** | - |
| Yes | 14 (9%) | 8 (3%) | 22 (5%) | - |  |  |
| No | 146 (91%) | 233 (92%) | 379 (92%) | - |  |  |
| Unknown | 0 | 13 (5%) | 13 (3%) | - |  |  |
| **Hospitalization** |  |  |  |  | **<0.001<sup>A</sup>** | - |
| Yes | 24 (15%) | 6 (2%) | 30 (7%) | - |  |  |
| No | 136 (85%) | 235 (93%) | 371 (90%) | - |  |  |
| Unknown | 0 | 13 (5%) | 13 (3%) | - |  |  |

<sup>A</sup> Wilcoxon test. <sup>B</sup> Fisher's exact test.
Post COVID-19 Condition (PCC); Inter Quartile Range (IQR); Body Mass Index (BMI); General Practitioner (GP).

Significant differences regarding sex, BMI, and age were observed in PCC patients compared to non-PCC cases, but these characteristics did not differ between the overall COVID-19 positive group (PCC and non-PCC cases combined) and community controls. This implies that our sampling method for community controls, aimed at a comparable distribution of main baseline characteristics between COVID-19 patients and community controls, was effective.

### Symptomatology

Among the SARS-CoV-2 positive cohort, presence of shortness of breath (OR=2.27 [1.07-4.94]) or reduced physical endurance (OR=3.34 [1.13-11.09]) prior to infection were significantly associated with PCC. All symptoms except chest pain were significantly more often experienced during the acute phase by patients that developed PCC than patients who did not (**Supplementary Table 6**). Among PCC patients, the most prevalent acute symptoms included fatigue (85%), reduced physical endurance (83%), headache (78%), and reduced muscle strength (76%) (**Supplementary Table 7**). Of non-PCC cases, 48% reported fatigue, 46% worsened physical endurance, 61% headache, and 39% reduced muscle strength in the acute phase.

In the post-acute phase, fatigue (67%) and reduced physical endurance (66%) were the most persisting symptoms. Persisting chest pain (37%) and sleeping problems (46%) were more prevalent among female PCC patients (vs. 22% and 28% among male patients, respectively). Common symptoms among patients over 60 years old included reduced physical endurance (79%), fatigue (76%) and loss of appetite (76%). Concentration problems, reduced physical endurance, shortness of breath, and reduced muscle strength were predominant among patients who had been hospitalized in the acute phase, as opposed to patients who had not been hospitalized (**Supplementary Table 7**). There was no significant association between presence of one or more comorbidities, sex, age group, or hospitalization during the acute phase and each of the fourteen symptoms in the post-acute phase (**Supplementary Table 7**).

### PCC screening based on a simple risk score

In total, 390 respondents were included in risk score analyses of which 40% (n =156) were PCC patients. The distributions of the risk scores are shown in **Figure 2** and the risk score formulas used to calculate a risk score per patient in **Supplementary Table 3**. Baseline characteristics for PCC patients and non-PCC cases included are shown in **Supplementary Table 8**. The risk score ranges calculated using the sparse risk score formulas are narrower compared to the risk score ranges calculated from the rich risk score formulas (**Figure 2**). The sparse risk scoring tool based on the full patient sample had an Area Under the Curve (AUC) of 0.68 (bootstrapped 95 CI% 0.63 – 0.74), whereas the sparse risk scoring tool based on the non-hospitalized patient sample had an AUC of 0.65 (bootstrapped 95% CI 0.59 – 0.71). The rich risk scoring tool based on the full patient sample had an AUC of 0.79 (95 CI% 0.74 – 0.83) and the rich risk scoring tool based on the non-hospitalized patient sample had an AUC of 0.77 (95CI% 0.72 -0.82). ROC curves are shown in **Supplementary Figure 3**. The graphs in **Figure 3** can be used to assess the trade-off between time and resource investment and identifying a greater number of people at risk of developing PCC. Generally, a higher risk score cut-off results in a higher PPV and lower NPV.

**Fig. 2.**
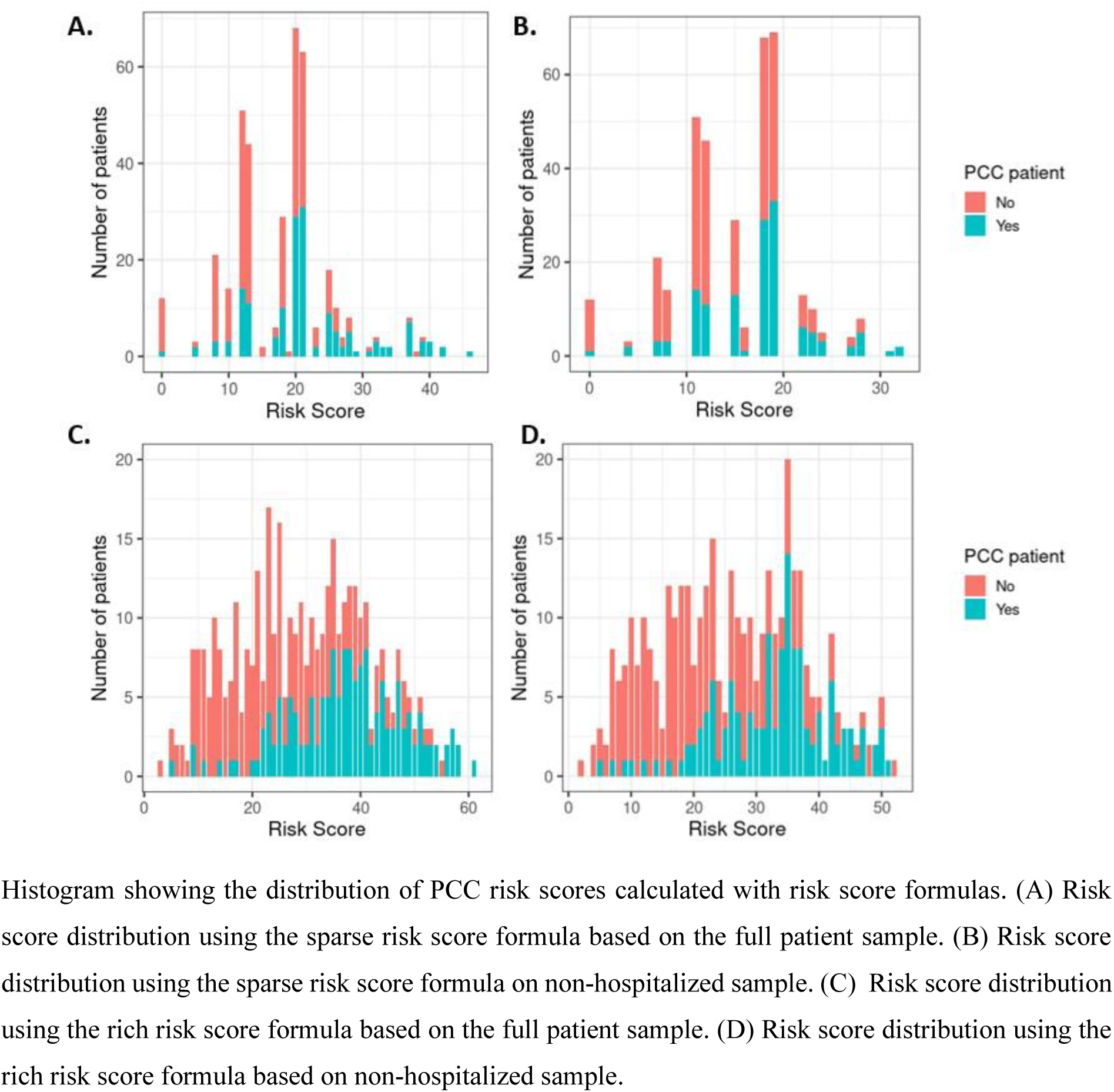
PCC risk score distributions

**Fig. 3.**
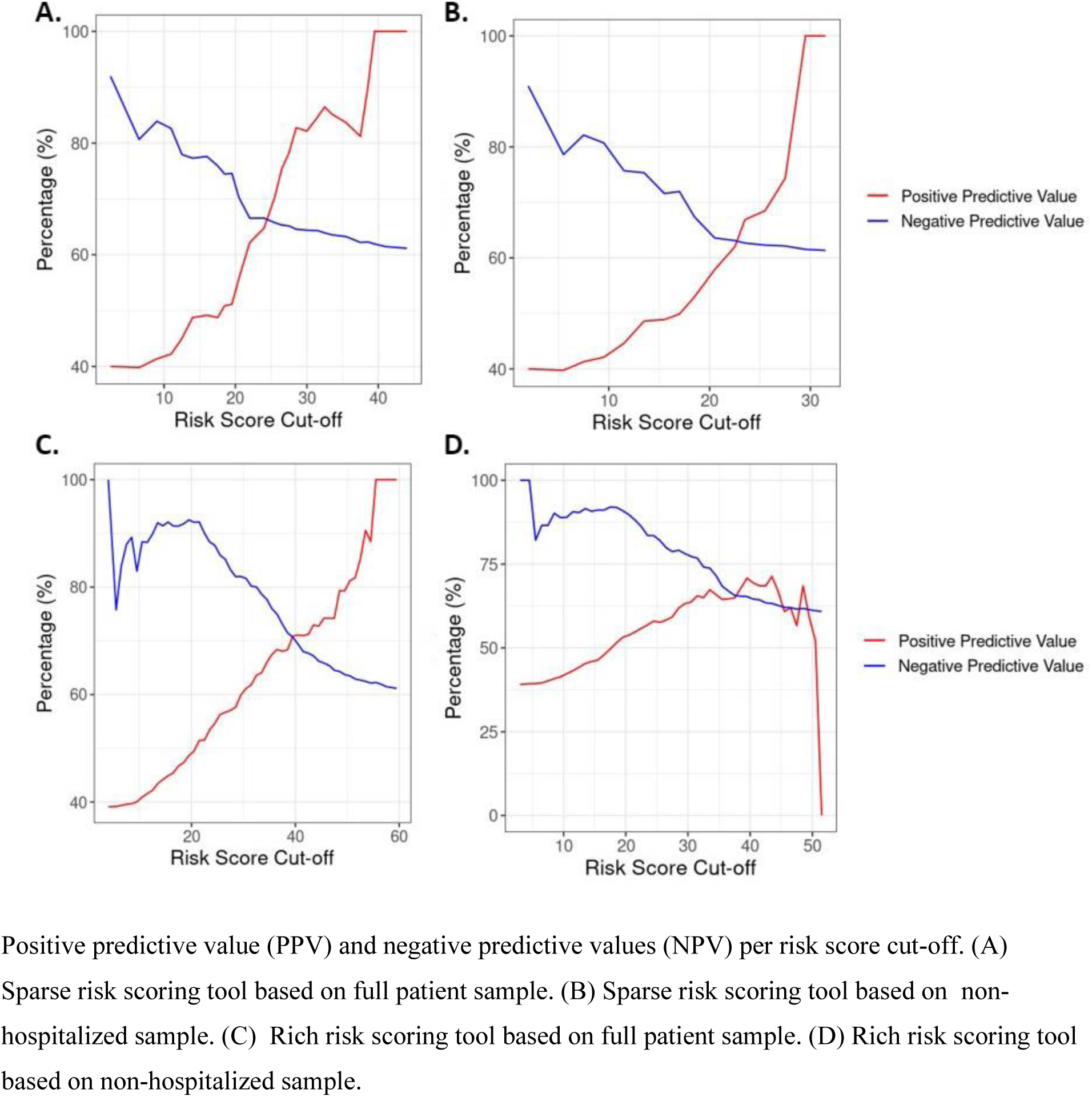
Positive and negative predictive values per risk score cut-off

### Worked example of using risk score for PCC screening

We provide worked examples for two scenarios in **Table 2**, using the graphs in **Figure 3**. The first scenario reflects a low-resource setting, where a high PPV and high risk score cut-off are desirable given the limited number of people that would have to be contacted. All those who would be contacted in this scenario would have a high probability of being true PCC patients. A disadvantage, however, is that in this scenario many other true PCC patients may miss out on being contacted. Contrarily, in the second, high resource scenario, a high NPV and low risk score cut-off may be desirable. Here, many patients would be contacted which would increase the number of false positives; however most true PCC patients would be reached.

**Table 2.** Number of patients who would be contacted (Call +) who truly have or do not have PCC for different risk score cut-offs

**A. Low-resource setting**
|  | PCC + | PCC - | Total |
| --- | --- | --- | --- |
| Hospitalized and non-hospitalized PCC patients<sup>a</sup> |  |  |  |
| Call + | 66 | 26 | 92 |
| Call - | 90 | 208 | 298 |
|  | 156 | 234 | 390 |
| Non-hospitalized PCC patients<sup>b</sup> |  |  |  |
| Call + | 64 | 36 | 100 |
| Call - | 70 | 192 | 262 |
|  | 134 | 228 | 362 |

<sup>a</sup> Risk score cut-off 40, using our rich risk scoring formula based on full patient sample.
<sup>b</sup> Risk score cut-off 35, using our rich risk scoring formula based on non-hospitalized sample.

**B. High resource setting**
|  | PCC + | PCC - | Total |
| --- | --- | --- | --- |
| Hospitalized and non-hospitalized PCC patients<sup>a</sup> |  |  |  |
| Call + | 151 | 183 | 334 |
| Call - | 5 | 51 | 56 |
|  | 156 | 234 | 390 |
| Non-hospitalized PCC patients<sup>b</sup> |  |  |  |
| Call + | 124 | 119 | 243 |
| Call - | 70 | 109 | 119 |
|  | 134 | 228 | 362 |

<sup>a</sup> Risk score cut-off 35, using our rich risk scoring formula based on full patient sample.
<sup>b</sup> Risk score cut-off 20, using our rich risk scoring formula based on non-hospitalized sample.

To illustrate the above points, we provide worked examples based on the rich risk scoring tool based on the full sample and the rich risk scoring tool based on the non-hospitalized sample for a low-and high-resource setting. Taking the low-resource setting as an example, our aim would be to identify as many true PCC patients whilst minimizing capacity going towards contacting efforts. Therefore, a high risk score cut-off and PPV is desirable; yet care must be taken not to choose a risk score that is at the tail end of the distribution, given few people will have such a score (**Figure 2**). Based on **Figure 3**, we chose a risk score of 40 with a PPV around 70%. Applying the risk scoring tool to our data, we found that 92 patients had a risk score of 40 or greater, of whom 66 (72%) were true PCC patients, yet 90 other true PCC patients would be missed as they would not be contacted.

### Sensitivity analysis

Our intermediate risk scoring tool for the full patient sample had an AUC of 0.71 (95% CI, 0.65 – 0.76) and the AUC was 0.67 (95% CI, 0.61-0.73) for the non-hospitalized patient sample (**Supplementary Table 5**).

## DISCUSSION

PCC is a debilitating condition and evidence regarding prevalence, symptomatology, and risk screening tools are lacking in the Caribbean context. In our study, 39% of COVID-19 cases on Bonaire experienced at least one persisting symptom for at least four weeks after disease onset. Most common persisting symptoms included fatigue (67%) and reduced physical endurance (66%).

Additionally, we describe proof-of-concept risk scoring tools using easily obtainable patient characteristics with good discriminatory power, which, after external validation in other study populations, could aid in the timely identification of those at increased risk of PCC. This is important because long-COVID rehabilitation care on Bonaire needs to be applied for within a specified time frame in order to be eligible for health insurance coverage [16].

### Prevalence

Symptom attribution to SARS-CoV-2 infection has shown to be challenging and there is substantial heterogeneity in prevalence estimates both within and between studies. This is caused by differences in case definition (such as duration of illness and type of symptoms) and differences in region, characteristics, and hospitalization status of the study populations [2, 6, 24]. For example, our estimate of 39% of symptomatic acute COVID-19 cases having symptoms for more than 4 weeks is in line with a recent global PCC pooled prevalence estimate of 43% (95% CI, 39-46) [2], however, it is lower than an estimate from a prospective cohort study in the European Netherlands of 49% of cases reporting at least one symptom three months after SARS-CoV-2 infection [6].

Our PCC case definition is based on the (CDC) working definition at the time of interview, using a cutoff of at least four weeks after disease onset [17]. The WHO case definition at the time of writing, on the other hand, indicates that a symptom should persist for at least 12 weeks after infection, of which at least 8 weeks cannot be explained by an alternative diagnosis [25]. This difference in follow-up time is important as its been shown to affect PCC prevalence estimates. Results of a meta-analysis analyzing global PCC estimates by Chen et al. (2022) reveal that PCC prevalence was 37 % when using a 30 day follow-up definition (95% CI, 26-49%), which decreased to 25% (95% CI, 15-38%) when a 60 day follow-up definition was used [2]. Surprisingly, prevalence across studies increased again to 32% (95% CI, 14-57%) and 49% (95% CI, 40-59%) using follow-up definitions of 90 and 120 days; possibly due to oversampling of hospitalization patients in the study populations of studies with longer follow up [2]. Using the WHO case definition, we observed a PCC prevalence 23% among our sample.

Consistent with other studies, we found that fatigue (67%) was the most common PCC symptom, followed by reduced physical endurance (66%) [26]. Among a sample of European Dutch and Belgian patients, Goertz et al. (2020) found fatigue (87%) and shortness of breath (71%) were most prevalent PCC symptoms [27]. Van der Maaden et al. (2023) found a significantly higher prevalence of fatigue (31.1%), dyspnea (16.4%), concentration problems (15.0%) and several other symptoms among cases from the European Netherlands as compared to control groups [6].

### Risk scoring tool

Besides gaining insight into PCC prevalence on Bonaire, practical and easy-to-use risk scoring tools for public and primary care are needed in low resource-settings to screen and identify persons that are at risk of developing PCC. Few PCC prediction tools exist and the ones that do, do not necessarily have practicality of use in mind – making them less usable for low-resource settings – nor are they applied to Caribbean populations. Our sparse risk scoring tool, using patient characteristics that are easily obtainable by GPs in practice (i.e. age, sex, presence of one or more comorbidities, hospitalization or GP visit for acute COVID-19); resulted in moderate discriminative power for the prediction of PCC. Adding smoking, alcohol use, BMI, education level, and number of symptoms in the acute phase improved the discriminative power considerably in the full patient sample (hospitalized plus non-hospitalized) as well as for the non-hospitalized patient sample.

Our results highlight the importance of obtaining certain patient characteristics for designing risk scoring tools either at intake or upon verbal inquiry with the patient. The moderate discriminatory performance of our sparse risk scoring tools – suggest that solely using baseline characteristic is not optimal in identifying patients at risk for PCC. Our rich risk scoring tools, on the other hand, showed a marked increase in discriminatory performance allowing for better delineation of PCC patients. Importantly, our risk scoring tools excluding hospitalized patients still obtained a good predictive performance aiding in the identification of PCC patients, which in absolute numbers is the largest patient population at risk for PPC. Sensitivity analysis showed a moderate performance of our intermediate risk scoring tool when including hospitalized patients. However, when excluding hospitalized patients, its discriminatory performance was poor-to-moderate and similar to our sparse risk scoring tool. This indicates that education level, and in particular the number of symptoms during the acute phase, are important predictors for PCC risk in the non-hospitalized population.

Although our rich risk scoring tools had a better discriminatory performance, calibration plots also show they overpredict PCC risk at higher predicted probabilities and underpredict at lower predicted probabilities (**Supplementary Figure 2**). This behavior is not necessarily problematic if risk scores are used as a PCC screening tool. In practice overoptimistic PCC risk prediction will result in a greater number of false positives. Although more false positives in this case would result in an increased use of capacity to contact patients, there would be negligible negative consequences for patients, as they could simply inform their GP whether they are experiencing persisting symptoms or not.

### Strengths & Limitations

We designed the first retrospective cohort study estimating the prevalence and characteristics for PCC in the Dutch Caribbean context and devised a simple risk scoring formula to estimate PCC risk. Our study adds valuable insight into PCC prevalence for the Caribbean region, which has been underrepresented in PCC research [15]. A major strength of our study was that we included a negative control group, age-sex matched to the COVID-19 cases, to assess baseline health status, and only few studies have done this thus far [24]. We found no significant difference in baseline demographic characteristics between the COVID-19 cohort and community controls, allowing an unbiased comparison of pre-existing health.

Contrary to most studies [24], we collected data on symptoms present before the SARS-CoV-2 infection for PCC patients, non-PCC cases, and community controls. Prior to the pandemic, symptoms were experienced to a similar or slightly higher degree by PCC patients than community controls, though we had limited statistical power to assess the significance of these differences. However, considering the vastly higher prevalence of symptoms post COVID-19 among PCC patients compared to their pre-pandemic experience and compared to the community controls, it is not likely that all PCC symptomatology can be explained as previously existing symptoms now being labeled as post-acute symptoms.

We did find that COVID-19 patients who reported already experiencing shortness of breath (OR=2.27 [1.07-4.94]) and reduced physical endurance (OR=3.34 [1.13-11.09]) prior to infection were more likely to suffer from post-acute symptoms than COVID-19 patients without these two pre-existing symptoms. A recent study using the European Netherlands Lifelines cohort data matched SARS-CoV-2 positive cases with a control group of SARS-CoV-2 negative participants and found that one in eight COVID-19 cases had persistent symptoms for 90-150 days after their acute COVID19 infection that could be attributed to their infection [2]. Our inclusive sampling strategy, i.e., inviting all residents on Bonaire who had received a SARS-CoV-2 positive test result before 1 October 2021 and experienced a symptomatic infection to participate in the study, supports the generalizability of our findings to the COVID-19 patient population on Bonaire.

As community controls were included based on self-reported SARS-CoV-2 negative status, PCC prevalence may be affected if these respondents were instead untested, SARS-CoV-2 positive cases. We opted to use patient reported data rather than using validated instruments for patient characteristics, previously registered patient data, or diagnostic tools for underlying disease, which would require more capacity and resources to obtain. Using self-reported data on presence of these symptoms may have led to information bias, though we believed our approach would result in more data considering the low-resource setting of our study. We did not expect potential information bias to impact our results, as we assumed this would not differ across subgroups. It is important to note that our risk score formula has not been externally validated. Our risk score formulas should therefore be interpreted as a proof-of-concept, and external validation would be needed before they can be utilized for PCC screening in general practice or public health settings.

### Implications for research, clinicians and policymakers

Our study adds valuable insight into PCC prevalence for the Caribbean region, which has been an underrepresented region for PCC research. Since the start of the pandemic, Bonaire reported 9,822 SARS-CoV-2 cases, of whom 98.5% concerned non-hospitalized cases with mild or asymptomatic infections (up to 8 January 2023) [9]. Applying our PCC prevalence of 39%, we expect that a substantial group of (former) COVID-19 patients may be experiencing persisting symptoms without seeking care at their GP, leaving the condition undiagnosed or with vast diagnostic delay among the at risk population [28]. GPs will require adequate tools to identify at risk patients, preferably using easy to use screening tools. Here, we highlight the added benefit of obtaining extra patient characteristics either at intake or upon verbal inquiry for improved PCC screening and stress the need for further research into external validation of risk scoring formulas.

## CONCLUSION

We estimated a PCC prevalence of 39% on Bonaire four weeks after disease onset. Moreover, easily obtainable patient characteristics can be used to build a risk scoring tool with moderate-to-good discriminatory power to identify those with PCC. If used for PCC screening, our risk scoring tool could aid public health in low-resource settings, however it needs to be externally validated first.

## Data Availability

All data produced in the present study are available upon reasonable request to the authors

## Acknowledgements

We would like to thank Adrienn Vegh (General Practitioner) who served as a practical advisor on post COVID-19 condition related primary care on Bonaire. Also, we would like to thank the team of interviewers from Tempo, Bonaire, who carried out the telephone interviews, and CBS Bonaire for their assistance in setting up training for the group of interviewers. Furthermore, we would like to thank Alicia Xuan-Krijgsman (Public Health Department Bonaire) for assistance during the preliminary phase of this study’s design and Karin Cox (Public Health Department Bonaire) for assisting in developing the call list of cases eligible for inclusion in the study.

## STATEMENTS & DECLARATIONS

### Funding

“This work was funded by the RIVM department CIb their RAC programme budget: Research budget (grant number 0113/2021, August 5th 2021)”.

### Competing Interests

The authors have no conflict of interest to declare that are relevant to the content of this article.

### Author Contributions

LJ secured funding for the performance of the study; LJ, DB, GM, IT and EG contributed to the study conception and design; LJ, GM and IT established data collection procedures; IT was responsible for data registry; DB and EG defined the statistical analysis plan; DB, TD, LJ, EG and GM contributed to the synthesis of key findings and interpretation of the results; DB performed the primary literature search, carried out descriptive and univariate analyses, and took co-lead in drafting and writing of the manuscript prior to publication. TD carried out descriptive analyses, quantitative analyses, and took co-lead in drafting the manuscript prior to publication; EG, LJ, IT and GM critically reviewed the manuscript.

### Ethics approval

The study protocol was sent to the Central Committee on Research Involving Human Subjects (CCMO Netherlands), who confirmed on 8 October 2021 that the study did not require ethical approval due to its observational design.

### Consent to participate

Informed consent was obtained from all individual participants included in the study.

### Consent to publish

Not applicable.

## Supplementary content

**Supplementary Table 1:** Variables used for model building

| Variable | Type | Description |
| --- | --- | --- |
| Age | Categorical | Age in years. Categories are: 0-19 years, 20-39 years, 40-59 years, and 60+ years old |
| Sex | Binary | Male, female. |
| Education level | Categorical | Categories are:<br>Low level education = no education, primary level education, lower vocational & lower secondary education, moderate level education = higher secondary education, high level education = university level & post-secondary education. |
| BMI | Categorical | BMI (kg/m<sup>2</sup>), categorized into underweight (<18), normal ($\geq 18$-<25), overweight ($\geq 25$-<30), obese ($\geq 30$-<35) and morbidly obese ($\geq 35$). |
| Smoking | Binary | Binary variable indicating whether or not a patient smoked cigarettes before the start of the pandemic. |
| Alcohol use | Binary | Binary variable indicating whether or not a patient used alcohol before the start of the pandemic. |
| Presence of one or more comorbidities | Binary | Binary variable indicating whether a person suffers from any of the following prior to the start of the pandemic: self-reported post-acute respiratory problems (including asthma, chronic obstructive pulmonary disorder, tuberculosis), post-acute cardiovascular disease, diabetes, severe kidney disease (including dialysis or kidney transplantation), HIV, severe liver disease, being immunocompromised, cancer, depression or anxiety, stroke, rheumatism, arthritis, or memory loss due to a neurological condition or dementia. |
| Number of symptoms in the acute phase | Numerical | Number of symptoms a patient experienced during the acute phase of their SARS-CoV-2 infection. Symptoms include: chest pain, concentration problems, cough, fatigue, headache, heart palpitations, loss of appetite, reduced muscle strength, loss of sense of smell, loss of sense of taste, muscle ache, reduced physical endurance, shortness of breath, sleeping problems. |
| GP visit in the acute phase | Binary | Binary variable indicating whether a patient visited or was visited by a GP during the acute phase of their SARS-CoV-2 infection. |
| Hospitalized during the acute phase | Binary | Binary variable indicating whether a patient was hospitalized during the acute phase of their SARS-CoV-2 infection. |

**Supplementary Table 2:** Coefficients from sparse and rich multivariate logistic regression models assessing the risk of PCC in full patient – and non-hospitalized patient samples

| Model | Age (ref cat: 0-19 years old) |  |  | Sex (ref: male) | Presence of one or more comorbidities (ref: no) | GP visit acute phase (ref: no) | Hospitalized during acute phase (ref: no) | Alcohol use (ref: no) | Smoking (ref: no) | BMI (ref: normal) |  |  |  | Education (ref: low) |  | Number of symptoms acute phase |
| --- | --- | --- | --- | --- | --- | --- | --- | --- | --- | --- | --- | --- | --- | --- | --- | --- |
|  | 20-39 | 40-60 | 60+ |  |  |  |  |  |  | Under weight | Over weight | Obese | Morbidly obese | Moderate | High |  |
| Sparse 1a | **1.17\*** | **1.27\*\*** | 1.02 | **0.80\*\*\*** | 0.47 | **0.66\*\*\*** | **1.91\*\*\*** | - | - | - | - | - | - | - | - | - |
| Sparse 1b | **1.12\*** | **1.16\*** | 0.75 | **0.72\*\*\*** | 0.43 | 0.90 | - | - | - | - | - | - | - | - | - | - |
| Rich 2a | 0.30 | 0.59 | 0.50 | 0.46 | 0.33 | 0.31 | **1.17\*** | 0.39 | -0.18 | 0.49 | 0.33 | 0.35 | **0.90\*\*** | 0.28 | 0.65 | **0.19\*\*\*** |
| Rich 2b | 0.23 | 0.46 | 0.14 | 0.37 | 0.23 | 0.59 | - | 0.37 | -0.06 | 0.44 | 0.23 | 0.37 | **0.90\*\*** | 0.31 | 0.63 | **0.19\*\*\*** |

**Note:** Reference categories are shown in each column. Significant coefficients are shown in bold; the Asterix refers to three categories: \*p<0.05, \*\*p<0.01, \*\*\*p<0.001. Models 1a and 2a use the full patient sample (n=390) and Models 1b and 2b use the non-hospitalized patient sample (n=362).

**Supplementary Table 3:** Risk score formulas to calculate the risk of PCC in COVID-19 patients

**Score table:**
| Age Category | Age Score |
| --- | --- |
| 0-19 | 0 |
| 20 – 39 | 12 |
| 40 – 60 | 13 |
| 60 + | 10 |

**Score table:**
| Age Category | Age Score |
| --- | --- |
| 0-19 | 0 |
| 20 – 39 | 11 |
| 40 – 60 | 12 |
| 60 + | 8 |

**Score table:**
| Age Category | Age Score |
| --- | --- |
| 0-19 | 0 |
| 20 – 39 | 3 |
| 40 – 60 | 6 |
| 60 + | 5 |

| **BMI Category** | **BMI Score** |
| --- | --- |
| Normal | 0 |
| Underweight | 5 |
| Overweight | 3 |
| Obese | 4 |
| Morbidly Obese | 9 |
| **Education Category** | **Education Score** |
| Low | 0 |
| Moderate | 3 |
| High | 7 |

**Score table:**
| **Age Category** | **Age Score** |
| --- | --- |
| 0-19 | 0 |
| 20 – 39 | 3 |
| 40 – 60 | 6 |
| 60 + | 5 |
| **BMI Category** | **BMI Score** |
| Normal | 0 |
| Underweight | 5 |
| Overweight | 3 |
| Obese | 4 |
| Morbidly Obese | 9 |
| **Education Category** | **Education Score** |
| Low | 0 |
| Moderate | 3 |
| High | 7 |

**Supplementary Table 4:**
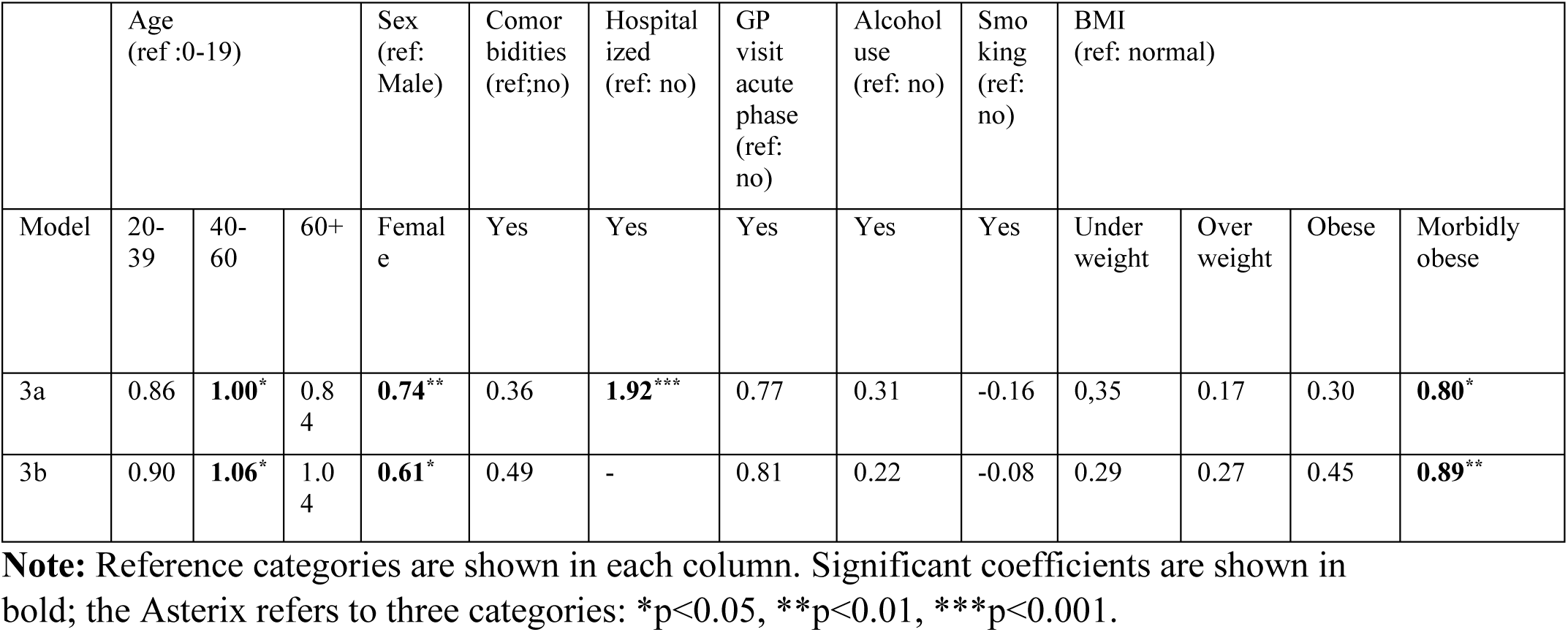
Coefficients from intermediate multivariate logistic regression models assessing the risk of PCC in full patient (3a)– and non-hospitalized patient (3b) samples

**Supplementary Table 5:** Sensitivity analyses for risk score formulas to calculate the risk of PCC in COVID-19 patients

| Age Category | Age Score |
| --- | --- |
| 0-19 | 0 |
| 20 – 39 | 9 |
| 40 – 60 | 10 |
| 60 + | 8 |
| BMI Category | BMI Score |
| Normal | 0 |
| Underweight | 4 |
| Overweight | 2 |
| Obese | 3 |
| Morbidly Obese | 8 |

| Age Category | Age Score |
| --- | --- |
| 0-19 | 0 |
| 20 – 39 | 9 |
| 40 – 60 | 11 |
| 60 + | 10 |
| BMI Category | BMI Score |
| Normal | 0 |
| Underweight | 3 |
| Overweight | 3 |
| Obese | 4 |
| Morbidly Obese | 9 |

**Supplementary Table 6:**
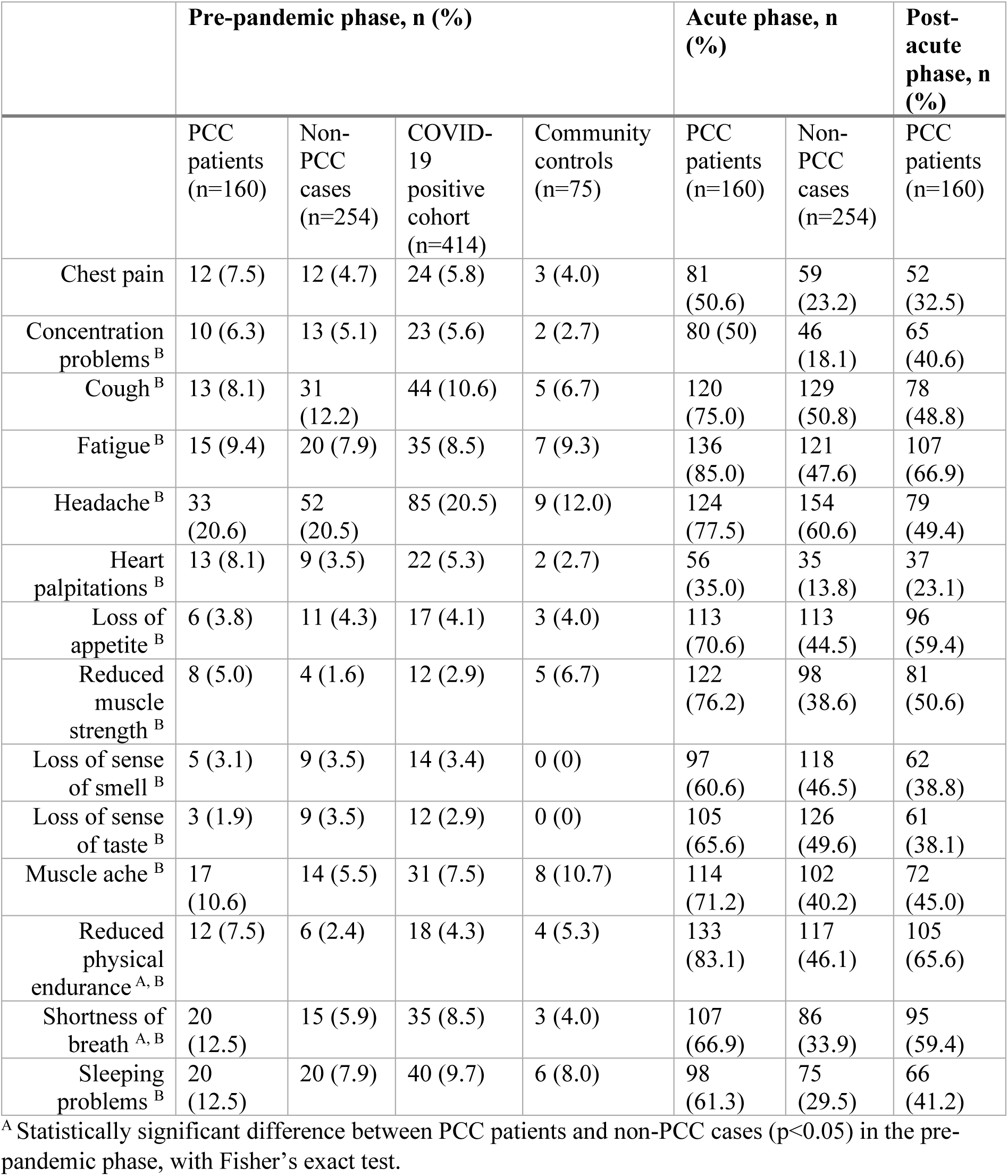
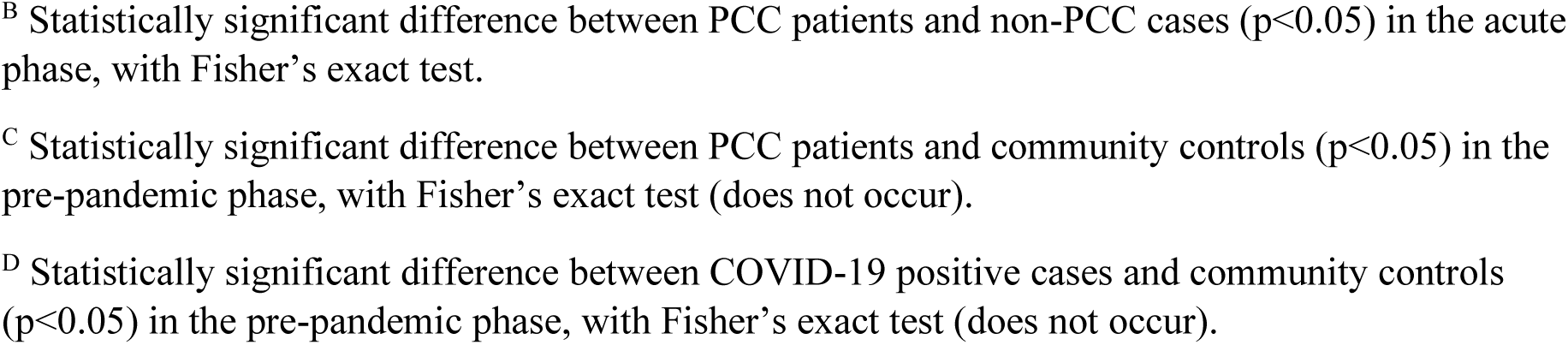
Prevalence of symptoms (n, %) by subgroup in the pre-pandemic, acute, and post-acute phase

|  | Pre-pandemic phase, n (%) |  |  |  | Acute phase, n (%) |  | Post-acute phase, n (%) |
| --- | --- | --- | --- | --- | --- | --- | --- |
|  | PCC patients (n=160) | Non-PCC cases (n=254) | COVID-19 positive cohort (n=414) | Community controls (n=75) | PCC patients (n=160) | Non-PCC cases (n=254) | PCC patients (n=160) |
| Chest pain | 12 (7.5) | 12 (4.7) | 24 (5.8) | 3 (4.0) | 81 (50.6) | 59 (23.2) | 52 (32.5) |
| Concentration problems<sup>B</sup> | 10 (6.3) | 13 (5.1) | 23 (5.6) | 2 (2.7) | 80 (50) | 46 (18.1) | 65 (40.6) |
| Cough<sup>B</sup> | 13 (8.1) | 31 (12.2) | 44 (10.6) | 5 (6.7) | 120 (75.0) | 129 (50.8) | 78 (48.8) |
| Fatigue<sup>B</sup> | 15 (9.4) | 20 (7.9) | 35 (8.5) | 7 (9.3) | 136 (85.0) | 121 (47.6) | 107 (66.9) |
| Headache<sup>B</sup> | 33 (20.6) | 52 (20.5) | 85 (20.5) | 9 (12.0) | 124 (77.5) | 154 (60.6) | 79 (49.4) |
| Heart palpitations<sup>B</sup> | 13 (8.1) | 9 (3.5) | 22 (5.3) | 2 (2.7) | 56 (35.0) | 35 (13.8) | 37 (23.1) |
| Loss of appetite<sup>B</sup> | 6 (3.8) | 11 (4.3) | 17 (4.1) | 3 (4.0) | 113 (70.6) | 113 (44.5) | 96 (59.4) |
| Reduced muscle strength<sup>B</sup> | 8 (5.0) | 4 (1.6) | 12 (2.9) | 5 (6.7) | 122 (76.2) | 98 (38.6) | 81 (50.6) |
| Loss of sense of smell<sup>B</sup> | 5 (3.1) | 9 (3.5) | 14 (3.4) | 0 (0) | 97 (60.6) | 118 (46.5) | 62 (38.8) |
| Loss of sense of taste<sup>B</sup> | 3 (1.9) | 9 (3.5) | 12 (2.9) | 0 (0) | 105 (65.6) | 126 (49.6) | 61 (38.1) |
| Muscle ache<sup>B</sup> | 17 (10.6) | 14 (5.5) | 31 (7.5) | 8 (10.7) | 114 (71.2) | 102 (40.2) | 72 (45.0) |
| Reduced physical endurance<sup>A, B</sup> | 12 (7.5) | 6 (2.4) | 18 (4.3) | 4 (5.3) | 133 (83.1) | 117 (46.1) | 105 (65.6) |
| Shortness of breath<sup>A, B</sup> | 20 (12.5) | 15 (5.9) | 35 (8.5) | 3 (4.0) | 107 (66.9) | 86 (33.9) | 95 (59.4) |
| Sleeping problems<sup>B</sup> | 20 (12.5) | 20 (7.9) | 40 (9.7) | 6 (8.0) | 98 (61.3) | 75 (29.5) | 66 (41.2) |

<sup>A</sup> Statistically significant difference between PCC patients and non-PCC cases ( $p < 0.05$ ) in the pre-pandemic phase, with Fisher's exact test.
<sup>B</sup> Statistically significant difference between PCC patients and non-PCC cases ( $p < 0.05$ ) in the acute phase, with Fisher's exact test.
<sup>C</sup> Statistically significant difference between PCC patients and community controls ( $p < 0.05$ ) in the pre-pandemic phase, with Fisher's exact test (does not occur).
<sup>D</sup> Statistically significant difference between COVID-19 positive cases and community controls ( $p < 0.05$ ) in the pre-pandemic phase, with Fisher's exact test (does not occur).

**Supplementary Table 7:**
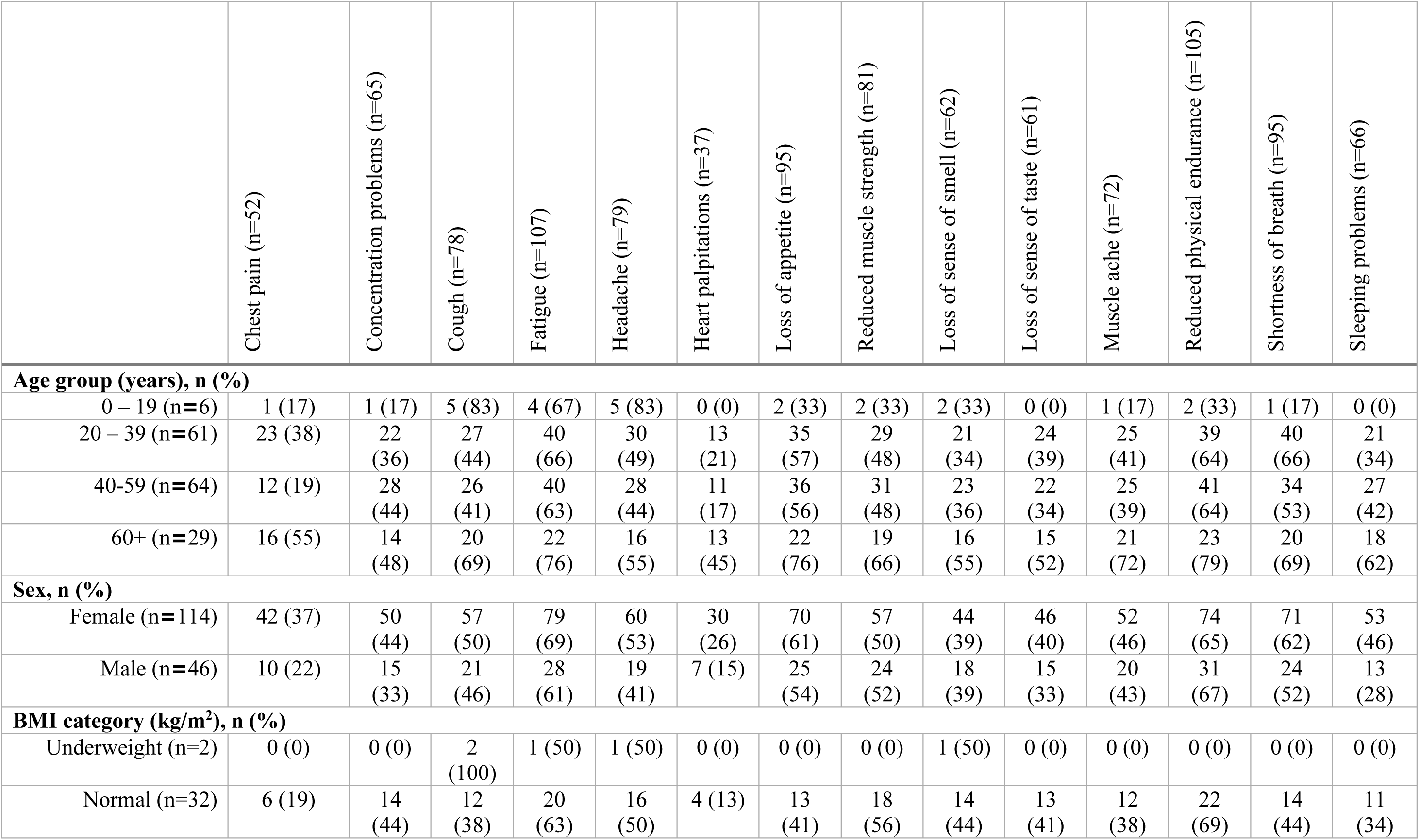
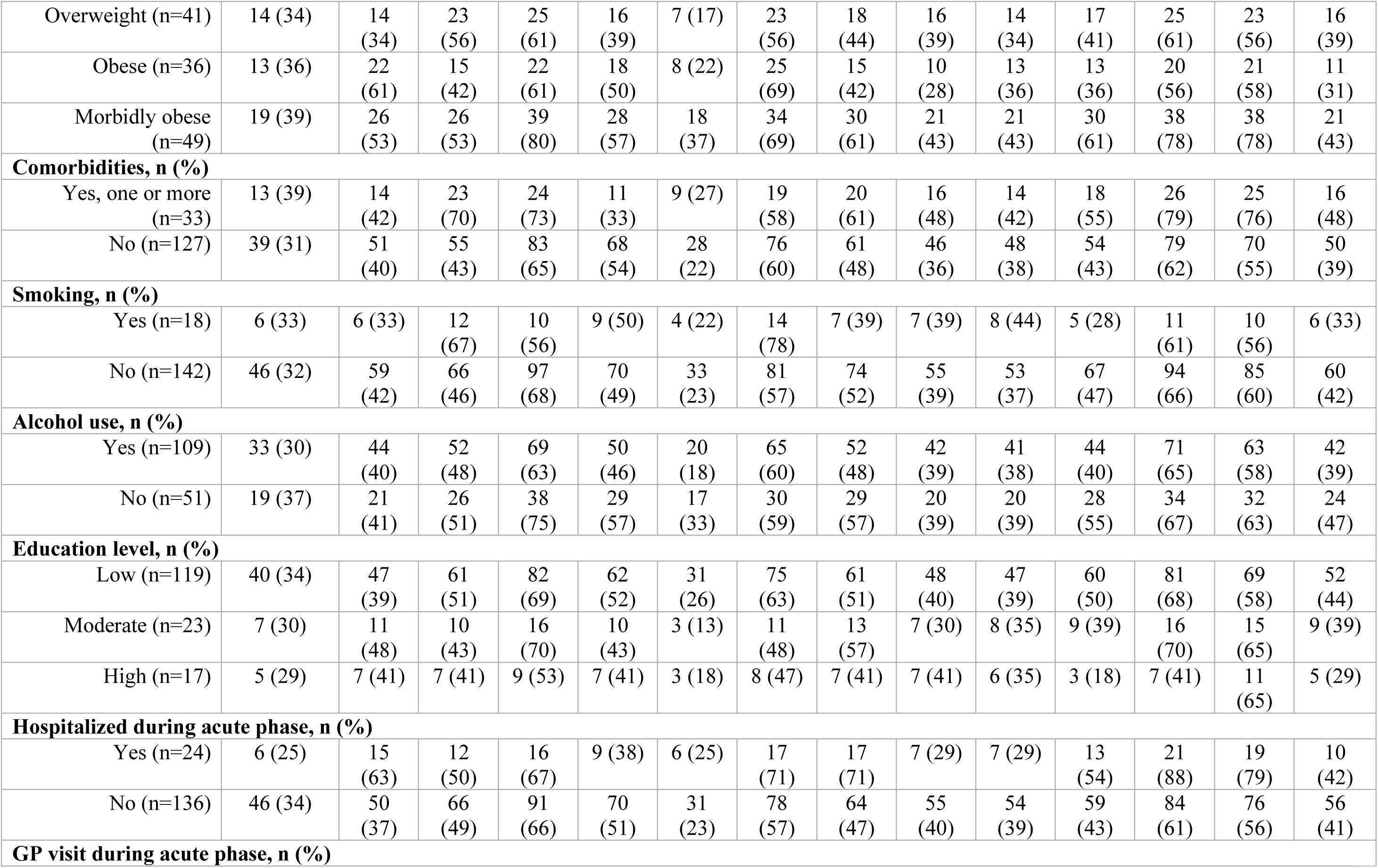
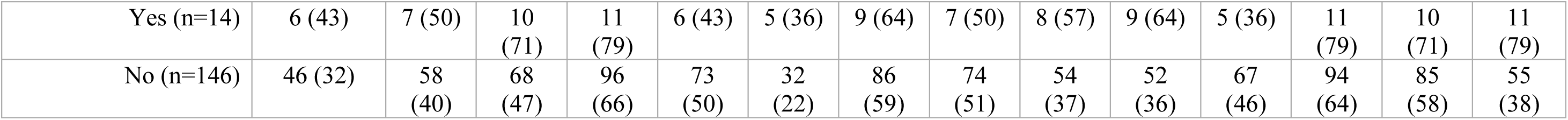
The prevalence of post-acute symptoms across different baseline characteristics of PCC patients (n=160)

|  | Chest pain (n=52) | Concentration problems (n=65) | Cough (n=78) | Fatigue (n=107) | Headache (n=79) | Heart palpitations (n=37) | Loss of appetite (n=95) | Reduced muscle strength (n=81) | Loss of sense of smell (n=62) | Loss of sense of taste (n=61) | Muscle ache (n=72) | Reduced physical endurance (n=105) | Shortness of breath (n=95) | Sleeping problems (n=66) |
| --- | --- | --- | --- | --- | --- | --- | --- | --- | --- | --- | --- | --- | --- | --- |
| **Age group (years), n (%)** |  |  |  |  |  |  |  |  |  |  |  |  |  |  |
| 0 – 19 (n=6) | 1 (17) | 1 (17) | 5 (83) | 4 (67) | 5 (83) | 0 (0) | 2 (33) | 2 (33) | 2 (33) | 0 (0) | 1 (17) | 2 (33) | 1 (17) | 0 (0) |
| 20 – 39 (n=61) | 23 (38) | 22 (36) | 27 (44) | 40 (66) | 30 (49) | 13 (21) | 35 (57) | 29 (48) | 21 (34) | 24 (39) | 25 (41) | 39 (64) | 40 (66) | 21 (34) |
| 40-59 (n=64) | 12 (19) | 28 (44) | 26 (41) | 40 (63) | 28 (44) | 11 (17) | 36 (56) | 31 (48) | 23 (36) | 22 (34) | 25 (39) | 41 (64) | 34 (53) | 27 (42) |
| 60+ (n=29) | 16 (55) | 14 (48) | 20 (69) | 22 (76) | 16 (55) | 13 (45) | 22 (76) | 19 (66) | 16 (55) | 15 (52) | 21 (72) | 23 (79) | 20 (69) | 18 (62) |
| **Sex, n (%)** |  |  |  |  |  |  |  |  |  |  |  |  |  |  |
| Female (n=114) | 42 (37) | 50 (44) | 57 (50) | 79 (69) | 60 (53) | 30 (26) | 70 (61) | 57 (50) | 44 (39) | 46 (40) | 52 (46) | 74 (65) | 71 (62) | 53 (46) |
| Male (n=46) | 10 (22) | 15 (33) | 21 (46) | 28 (61) | 19 (41) | 7 (15) | 25 (54) | 24 (52) | 18 (39) | 15 (33) | 20 (43) | 31 (67) | 24 (52) | 13 (28) |
| **BMI category (kg/m<sup>2</sup>), n (%)** |  |  |  |  |  |  |  |  |  |  |  |  |  |  |
| Underweight (n=2) | 0 (0) | 0 (0) | 2 (100) | 1 (50) | 1 (50) | 0 (0) | 0 (0) | 0 (0) | 1 (50) | 0 (0) | 0 (0) | 0 (0) | 0 (0) | 0 (0) |
| Normal (n=32) | 6 (19) | 14 (44) | 12 (38) | 20 (63) | 16 (50) | 4 (13) | 13 (41) | 18 (56) | 14 (44) | 13 (41) | 12 (38) | 22 (69) | 14 (44) | 11 (34) |
| Overweight (n=41) | 14 (34) | 14 (34) | 23 (56) | 25 (61) | 16 (39) | 7 (17) | 23 (56) | 18 (44) | 16 (39) | 14 (34) | 17 (41) | 25 (61) | 23 (56) | 16 (39) |
| Obese (n=36) | 13 (36) | 22 (61) | 15 (42) | 22 (61) | 18 (50) | 8 (22) | 25 (69) | 15 (42) | 10 (28) | 13 (36) | 13 (36) | 20 (56) | 21 (58) | 11 (31) |
| Morbidly obese (n=49) | 19 (39) | 26 (53) | 26 (53) | 39 (80) | 28 (57) | 18 (37) | 34 (69) | 30 (61) | 21 (43) | 21 (43) | 30 (61) | 38 (78) | 38 (78) | 21 (43) |
| **Comorbidities, n (%)** |  |  |  |  |  |  |  |  |  |  |  |  |  |  |
| Yes, one or more (n=33) | 13 (39) | 14 (42) | 23 (70) | 24 (73) | 11 (33) | 9 (27) | 19 (58) | 20 (61) | 16 (48) | 14 (42) | 18 (55) | 26 (79) | 25 (76) | 16 (48) |
| No (n=127) | 39 (31) | 51 (40) | 55 (43) | 83 (65) | 68 (54) | 28 (22) | 76 (60) | 61 (48) | 46 (36) | 48 (38) | 54 (43) | 79 (62) | 70 (55) | 50 (39) |
| **Smoking, n (%)** |  |  |  |  |  |  |  |  |  |  |  |  |  |  |
| Yes (n=18) | 6 (33) | 6 (33) | 12 (67) | 10 (56) | 9 (50) | 4 (22) | 14 (78) | 7 (39) | 7 (39) | 8 (44) | 5 (28) | 11 (61) | 10 (56) | 6 (33) |
| No (n=142) | 46 (32) | 59 (42) | 66 (46) | 97 (68) | 70 (49) | 33 (23) | 81 (57) | 74 (52) | 55 (39) | 53 (37) | 67 (47) | 94 (66) | 85 (60) | 60 (42) |
| **Alcohol use, n (%)** |  |  |  |  |  |  |  |  |  |  |  |  |  |  |
| Yes (n=109) | 33 (30) | 44 (40) | 52 (48) | 69 (63) | 50 (46) | 20 (18) | 65 (60) | 52 (48) | 42 (39) | 41 (38) | 44 (40) | 71 (65) | 63 (58) | 42 (39) |
| No (n=51) | 19 (37) | 21 (41) | 26 (51) | 38 (75) | 29 (57) | 17 (33) | 30 (59) | 29 (57) | 20 (39) | 20 (39) | 28 (55) | 34 (67) | 32 (63) | 24 (47) |
| **Education level, n (%)** |  |  |  |  |  |  |  |  |  |  |  |  |  |  |
| Low (n=119) | 40 (34) | 47 (39) | 61 (51) | 82 (69) | 62 (52) | 31 (26) | 75 (63) | 61 (51) | 48 (40) | 47 (39) | 60 (50) | 81 (68) | 69 (58) | 52 (44) |
| Moderate (n=23) | 7 (30) | 11 (48) | 10 (43) | 16 (70) | 10 (43) | 3 (13) | 11 (48) | 13 (57) | 7 (30) | 8 (35) | 9 (39) | 16 (70) | 15 (65) | 9 (39) |
| High (n=17) | 5 (29) | 7 (41) | 7 (41) | 9 (53) | 7 (41) | 3 (18) | 8 (47) | 7 (41) | 7 (41) | 6 (35) | 3 (18) | 7 (41) | 11 (65) | 5 (29) |
| **Hospitalized during acute phase, n (%)** |  |  |  |  |  |  |  |  |  |  |  |  |  |  |
| Yes (n=24) | 6 (25) | 15 (63) | 12 (50) | 16 (67) | 9 (38) | 6 (25) | 17 (71) | 17 (71) | 7 (29) | 7 (29) | 13 (54) | 21 (88) | 19 (79) | 10 (42) |
| No (n=136) | 46 (34) | 50 (37) | 66 (49) | 91 (66) | 70 (51) | 31 (23) | 78 (57) | 64 (47) | 55 (40) | 54 (39) | 59 (43) | 84 (61) | 76 (56) | 56 (41) |
| **GP visit during acute phase, n (%)** |  |  |  |  |  |  |  |  |  |  |  |  |  |  |
| Yes (n=14) | 6 (43) | 7 (50) | 10<br>(71) | 11<br>(79) | 6 (43) | 5 (36) | 9 (64) | 7 (50) | 8 (57) | 9 (64) | 5 (36) | 11<br>(79) | 10<br>(71) | 11<br>(79) |
| No (n=146) | 46 (32) | 58<br>(40) | 68<br>(47) | 96<br>(66) | 73<br>(50) | 32<br>(22) | 86<br>(59) | 74<br>(51) | 54<br>(37) | 52<br>(36) | 67<br>(46) | 94<br>(64) | 85<br>(58) | 55<br>(38) |

**Supplementary Table 8:** Characteristics of SARS-CoV-2 positive cases (n= 390) included in risk scoring tool analyses

|  | **PCC patients (n=156)** | **Non-PCC cases (n=234)** | **P-value** |
| --- | --- | --- | --- |
| **Age group (years), n (%)** |  |  |  |
| 0-19 years | 6 (34%) | 31 (13%) | **0.01<sup>B</sup>** |
| 20 – 39 years | 61 (39%) | 90 (39%) |  |
| 40 – 59 years | 63 (40%) | 80 (34%) |  |
| 60 + years | 26 (17%) | 33 (14%) |  |
| **Sex, n (%)** |  |  | **<0.01<sup>B</sup>** |
| Male | 45 (29%) | 107 (46%) |  |
| Female | 111 (71%) | 127 (54%) |  |
| **Comorbidities, n (%)** |  |  | **0.04<sup>B</sup>** |
| Yes | 30 (19%) | 27 (12%) |  |
| No | 126 (81%) | 207 (89%) |  |
| **GP visit in acute phase, n (%)** |  |  | **0.02<sup>B</sup>** |
| Yes | 14 (9%) | 8 (3%) |  |
| No | 142 (91%) | 226 (97%) |  |
| **Hospitalized for COVID-19 in acute phase, n (%)** |  |  | **<0.01<sup>B</sup>** |
| Yes | 22 (14%) | 6 (23%) |  |
| No | 134 (86%) | 228 (97%) |  |
| **Median number of symptoms in acute phase (IQR)** | 10 (5) | 5 (7) | **<0.01<sup>A</sup>** |
| **Pre-pandemic alcohol use** |  |  | **0.81<sup>B</sup>** |
| Yes | 106 (68%) | 148 (63%) |  |
| No | 50 (32%) | 86 (37%) |  |
| **Pre-pandemic smoking** |  |  | **0.63<sup>B</sup>** |
| Yes | 18 (12%) | 32 (14%) |  |
| No | 138 (89%) | 202 (86%) |  |
| **BMI (kg/m<sup>2</sup>) category** |  |  | **<0.01<sup>B</sup>** |
| Underweight | 2 (1%) | 4 (2%) |  |
| Normal | 32 (21%) | 75 (32%) |  |
| Overweight | 40 (26%) | 70 (30%) |  |
| Obese | 35 (22%) | 48 (21%) |  |
| Morbidly obese | 47 (30%) | 37 (16%) |  |
| **Education level** |  |  | **0.08<sup>B</sup>** |
| Low | 116 (74%) | 194 (83%) |  |
| Moderate | 24 (15%) | 27 (12%) |  |
| High | 17 (11%) | 13 (6%) |  |

<sup>A</sup> Wilcoxon test. <sup>B</sup> Fisher's exact test.

**Supplementary Figure 1:**
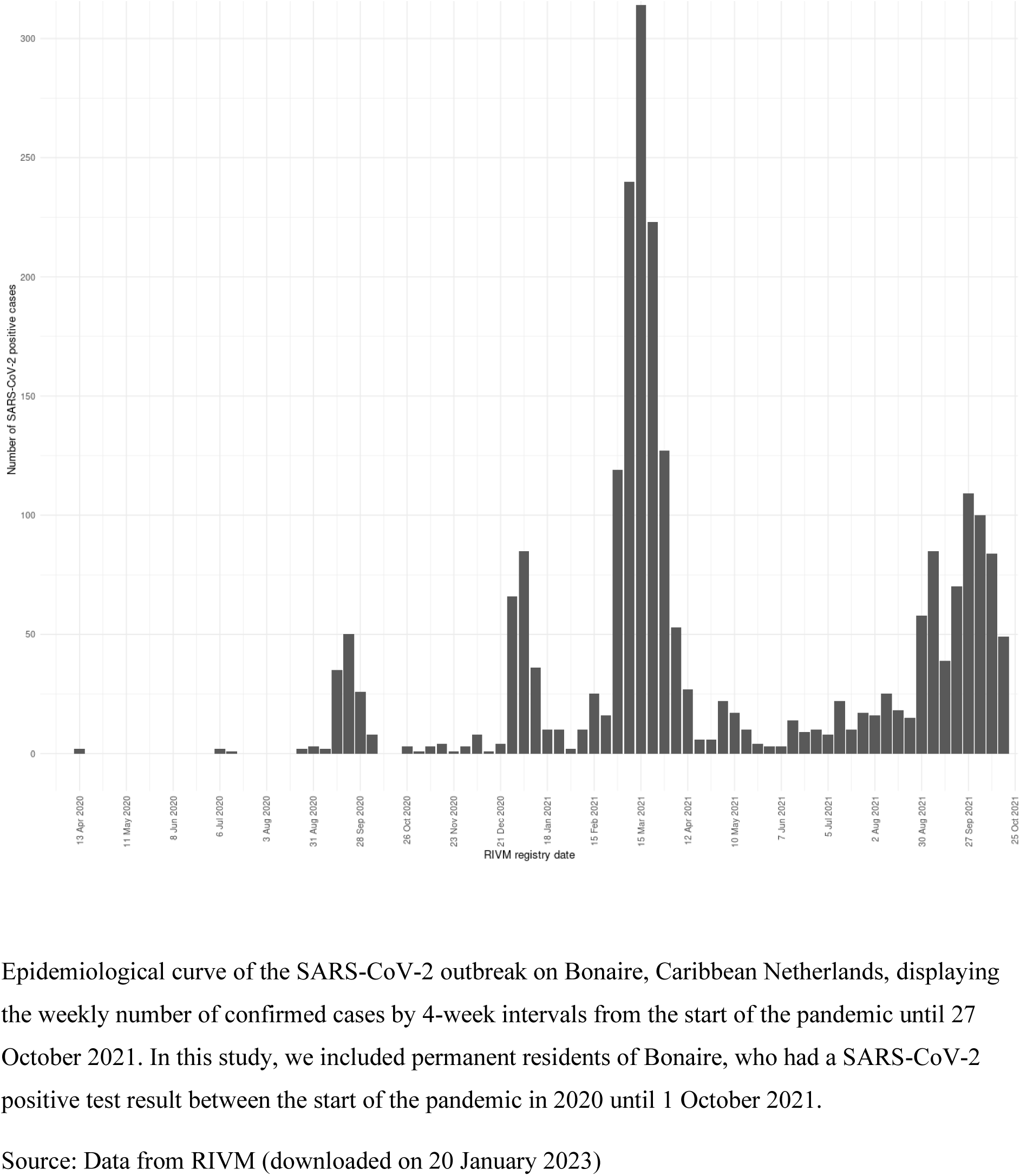
Epiurve and inclusion period for SARS-CoV-2 positive cases on Bonaire, Caribbean Netherlands

**Supplementary Figure 2:**
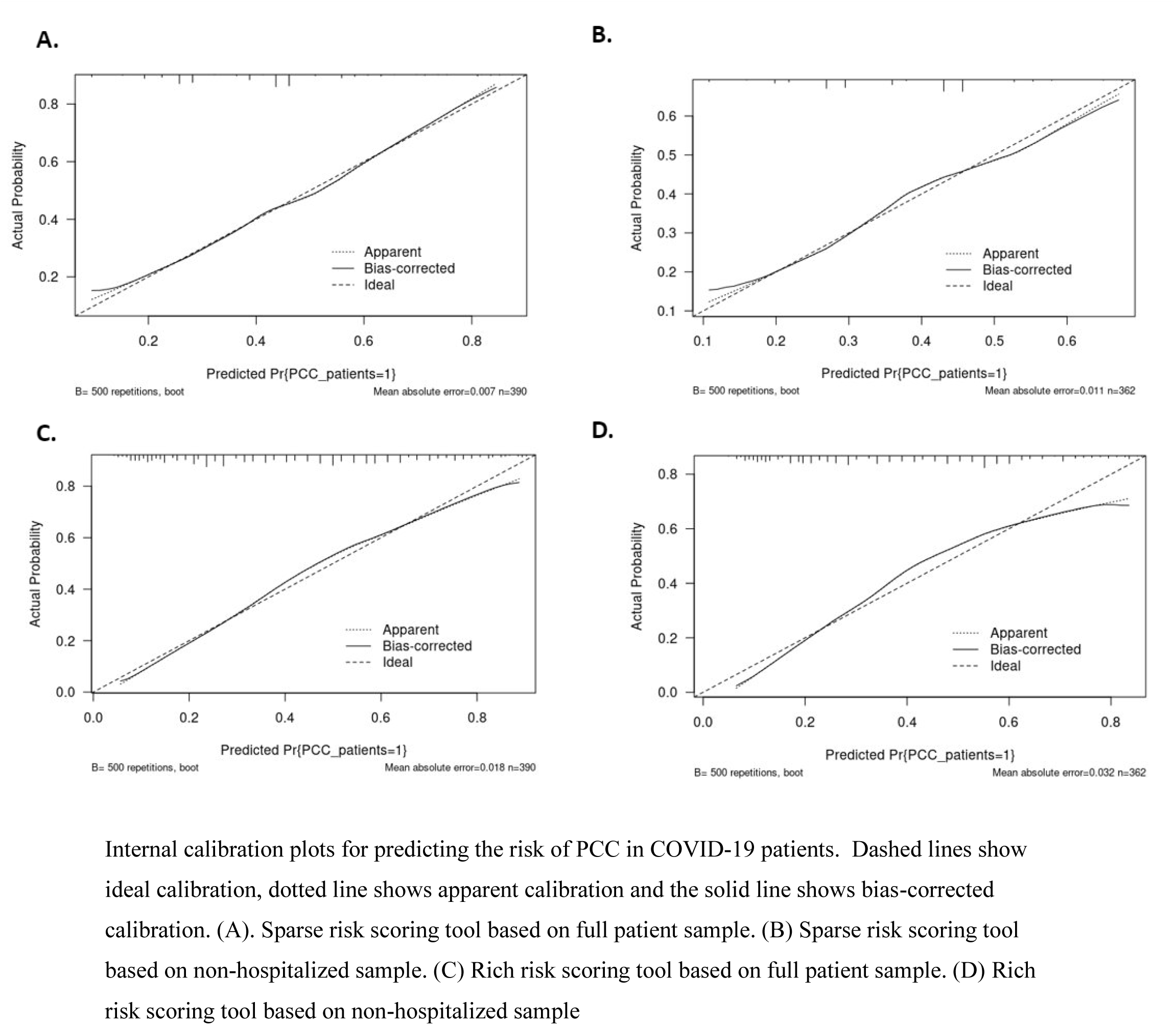
Internal calibration plots for predicting the risk of PCC PCC in COVID-19 patients

**Supplementary Figure 3:**
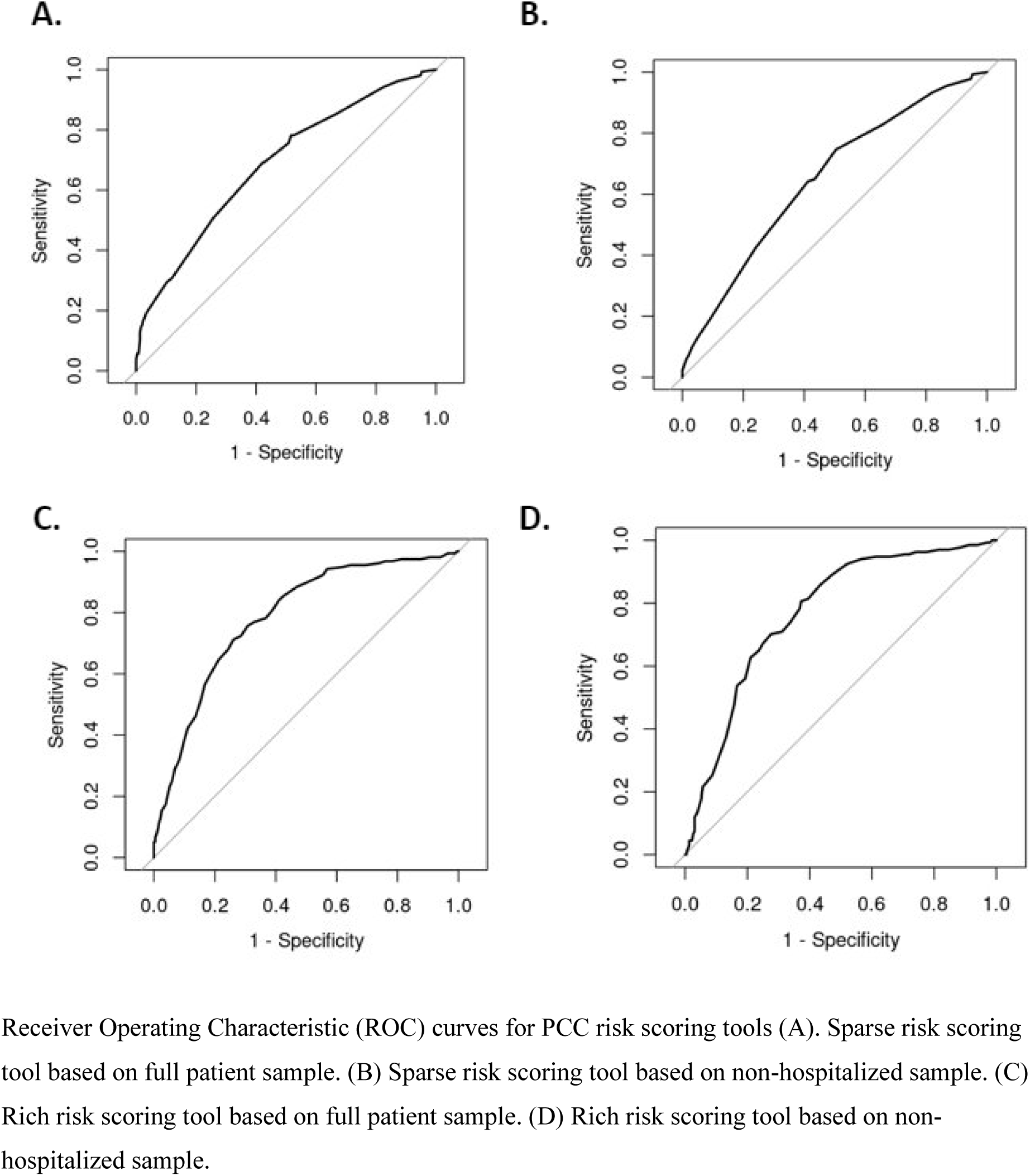
Receiver Operating Characteristic (ROC) curves for PCC risk scoring tools

## Notes

### Competing Interest Statement

The authors have declared no competing interest.

